# Association of Deep Learning-Derived Temporalis Sarcopenia with Mortality in Acute Ischemic Stroke

**DOI:** 10.64898/2026.01.21.26344571

**Authors:** Joshua Ron Onichino, Ding Yi Zhang, Miguel Trottier, Lorne Rosenbloom, Jonathan Afilalo

**Author notes:** **Corresponding Author:** Dr. Jonathan Afilalo, MD, MSc, FACC Director, Geriatric Cardiology Program, McGill University Division of Cardiology and Centre for Clinical Epidemiology, Jewish General Hospital 3755 Cote Ste Catherine, H-411, Montreal, Québec, Canada, H3T 1E2 |.

## Abstract

**Background:** Sarcopenia is associated with mortality and morbidity following acute ischemic stroke (AIS), but the diagnosis requires specialized equipment or time-consuming assessments. Computed tomography (CT) measures of temporal muscle volume (TMV) and density (TMD) can be opportunistically measured from existing scans and automated using deep learning (DL). This study sought to demonstrate the incremental prognostic value of DL-derived TMV and TMD from CT scans on mortality and length of stay (LOS) in AIS.

**Methods:** In this retrospective, single-centre cohort study, consecutive AIS patients admitted from 2014 to 2023 were included. Admission CT scans were retrieved alongside clinical data from electronic health records. TMV and TMD were quantified by a novel DL model and represented as continuous or trichotomous categorical variables. TMV and TMD thresholds were derived in a cohort of 50 healthy adults and used to classify AIS patients as non-sarcopenic, pre-sarcopenic, or sarcopenic. The primary outcome was 30-day all-cause mortality. Secondary outcomes were 365-day all-cause mortality and LOS. Multivariable logistic and linear regression were used.

**Results:** The cohort consisted of 2285 patients with 1151 (50%) females, and a mean (SD) age of 74.7 (13.7) years. Based on TMV and TMD, 877 patients (38%) were non-sarcopenic, 838 (37%) pre-sarcopenic, and 570 (25%) sarcopenic. Adjusted ORs for 30-day mortality were 2.70 (1.64 to 4.46) and 2.91 (1.72 to 4.91) for pre-sarcopenia and sarcopenia. Adjusted ORs for 365-day mortality were 2.42 (95% CI 1.74 to 3.36) and 2.96 (95% CI 2.09 to 4.17) and the additional days in hospital were 2.79 (1.69 to 3.98) and 3.26 (2.00 to 4.64) for pre-sarcopenia and sarcopenia. The association between CT-derived sarcopenia and mortality was preserved after adding HFRS to the models.

**Conclusions:** TMV and TMD extracted using a novel DL model were incrementally predictive of AIS mortality. These metrics may be used to refine risk estimates, inform shared decision-making, and individualize treatment plans.

## INTRODUCTION

Acute ischemic stroke (AIS) is a leading cause of death and disability worldwide.^1^ It constitutes two-thirds of incident strokes, resulting in over 7.8 million new cases annually^1^ and its incidence is rising in tandem with population aging.^1,2^ Age is associated with higher mortality and disability after AIS,^3–5^ but older patients have heterogeneous levels of functioning even after considering their comorbidities. Frailty, a state of diminished physiologic reserve,^6^ captures vulnerability beyond chronological age and comorbidity, and is increasingly used for risk stratification.^7^

Sarcopenia, a core component of frailty that is defined as age-related loss of muscle mass and function,^8^ is diagnosed in 46% of patients with stroke ^9^ and is associated with poor functional outcomes.^10^ However, it remains challenging to assess sarcopenia in clinical settings because assessment tools are often not reliable or feasible due to paresis, aphasia, or altered consciousness.^11–13^

Imaging biomarkers offer a practical alternative to conventional sarcopenia tests. Temporal muscle size and density can be measured from routine computed tomography (CT) imaging of the head. Emerging research suggests that reductions in temporal muscle thickness (TMT) and area (TMA) are associated with poor functional outcome and mortality in small cohorts of patients with ischemic^14,15^ and hemorrhagic stroke,^16,17^ although results have varied.^18,19^

Sources of inconsistency include reliance on manual measurements of one- or two-dimensional temporal muscle size (as opposed to three-dimensional volume) and omission of density as a biomarker of temporal muscle quality. While temporal muscle volume (TMV) reflects muscle mass, temporal muscle density (TMD) reflects intramuscular fat infiltration and thus muscle quality.^20^ General sarcopenia guidelines emphasize the need to consider both muscle mass and either function or quality, since low muscle mass alone is insufficient to diagnose sarcopenia.^8^

Both TMV and TMD arise from shared catabolic pathways that link sarcopenia to systemic inflammation and metabolic dysregulation, which plausibly worsen cerebral injury.^21^ To our knowledge, three-dimensional metrics of temporal muscle size and density have yet to be evaluated in AIS. Recent advances in deep learning (DL) now permit rapid, automated segmentation of CT scans, eliminating intra- and inter-observer variability and rendering large-scale implementation feasible.^22^

The objective of this study was twofold: first, to develop a novel DL model to automate TMV and TMD measurement from clinical CT scans, and second, to determine whether these metrics are incrementally predictive of short- and mid-term mortality and hospital length of stay (LOS) in AIS patients.

## METHODS

### Study Design and Setting

Consecutive AIS patients admitted to the Jewish General Hospital (JGH), a tertiary care centre in Montreal, Quebec, Canada, were retrospectively included in this single-centre cohort study between January 1, 2014, and December 31, 2023. Admission CT scans for each patient were retrieved alongside diagnostic data and vital status from electronic health records. Clinical CT scans were acquired on two multi-detector scanners, PrizMed GE Lightspeed VCT (64-slice) and Siemens SOMATOM Edge (128-slice), following the institutional stroke protocol. Axial images were reconstructed at 3-mm thickness (in-plane) with a soft-tissue reconstruction kernel. Exclusion criteria were confounding diagnoses, artifacts that rendered the scan uninterpretable, or missing CT images. The study was approved by the CIUSSS West-Central Montreal Research Ethics Board, which granted a waiver of individual informed consent for secondary analyses of existing clinical and imaging data. Requests for data sharing will be considered on a case-by-case basis following institutional governance policies.

### Temporal Muscle Segmentation

Manual segmentation was performed using 3D Slicer version 5.6.2 with a smart paintbrush tool configured to include voxels within a predefined intensity range of 25-95 Hounsfield units (HU) to isolate muscle tissue. The entire temporal muscle was segmented bilaterally on contiguous axial slices from its mandibular insertion on the coronoid process to its cranial origin along the temporal fossa.

### Deep Learning Model

Model training was conducted on clinical CT scans from the JGH and Dossier Santé Québec using a three-round, iterative nnU-Net protocol^22^ that progressively expanded the training set from an initial 40 scans (Figure S1). In Round 1, initial scans were manually segmented by a trained physician supervised by a board-certified neuroradiologist to generate a labelled training set and train an initial model. In Round 2, this model was applied to segment an additional 30 scans, which were manually corrected and incorporated into the training set to train an improved model. In Round 3, this model was applied to segment an additional 100 scans, again manually corrected, yielding a final training set of 170 labelled scans used to train the final DL model that met the performance benchmarks to obviate further rounds. The target performance benchmark was a Dice-Sorensen Coefficient (DSC) > 95% in the validation set. After iterative training, the final model was evaluated on a hold-out test set of 30 labelled scans, achieving a DSC of 94%.

### Temporal Muscle Metrics

The DL model was used to generate temporal muscle segmentations from the CT scans from which TMV (in cm³) and TMD (in HU) were extracted. TMV was the combined volume of both temporal muscles while TMD was the mean density across both temporal muscle volumes. To index measurements of TMV for head size, another DL model^23^ was used to generate skull segmentations. These segmentations were used to extract total skull volume (in cm^3^). Normative thresholds were established from a reference cohort of 50 clinically healthy adults with Charlson Comorbidity Index scores of 0 and no history of cerebrovascular disease, vascular risk factors, or inflammatory conditions. 5th percentile bootstrap quantile regression was used to define the lower reference value for TMV and TMD. Low TMV was 63.8 cm^3^ in males and 30.2 cm^3^ in females. Low TMD was 52.8 HU for both sexes.

Based on these normative thresholds, patients were classified as non-sarcopenic (normal TMV and normal TMD), pre-sarcopenic (low TMV or low TMD), or sarcopenic (low TMV and low TMD).

### Covariates

Baseline clinical risk was summarized by a validated in-hospital ischemic stroke mortality score^3^ that assigns weighted points to age, mode of arrival, arrival during regular hours, and nine pre-existing conditions (atrial fibrillation, prior stroke or transient ischemic attack, coronary artery disease, carotid stenosis > 50%, diabetes mellitus, peripheral vascular disease, hypertension, history of dyslipidemia, smoker). The total score returned a continuous predicted mortality probability that was treated as a single composite covariate.

Additionally, TMV and TMD were compared against frailty, measured by the Hospital Frailty Risk Score (HFRS),^24^ to assess whether CT-derived sarcopenia provides prognostic information independent of a validated frailty score. Continuous HFRS was entered when modelling continuous predictors (TMV and TMD), whereas categorical HFRS, in which patients were classified as low (< 5), intermediate (5-15), or high (> 15) risk, was entered when modelling categorical predictors (sarcopenia status).

### Outcomes

The primary outcome was 30-day all-cause mortality. Secondary outcomes were 365-day all-cause mortality and LOS. Mortality was determined from institutional electronic health records and confirmed by linked administrative data. LOS was defined as number of calendar days from hospital arrival to discharge. Given the skewed distribution of LOS, log-transformed LOS was used for modelling purposes.

### Statistical Analysis and Clinical Prediction Models

Continuous variables were reported as mean (SD), and categorical variables as number (%). Logistic regression was used to assess associations between temporal muscle metrics and mortality. Linear regression was used to assess associations between temporal muscle metrics and log-transformed LOS, which was transformed back into days for reporting. Model performance was assessed by area under the receiver operating characteristic (AUROC) curve analysis and residual diagnostics. Statistical analyses were performed using Stata version 19 software (StataCorp LLC). Two-sided *P* values < 0.05 were considered statistically significant. Dr. Jonathan Afilalo had full access to all the data and takes responsibility for the integrity of the data and the accuracy of the analyses.

## RESULTS

### Study Flow and Patient Characteristics

Figure S2 shows the flow diagram from initial to final cohort after applying exclusion criteria. The final cohort consisted of 2285 patients with 1151 (50%) females and a mean (SD) age of 74.7 (13.7) years. Excluded patients demonstrated similar characteristics. Mean (SD) TMV was 70.2 (24.9) cm^3^ in males and 43.1 (17.0) cm^3^ in females (Figure S3A). Mean (SD) TMD was 51.8 (5.1) HU in males and females (Figure S3B). Figure 1 shows representative CT scan segmentations.

**Figure 1.**
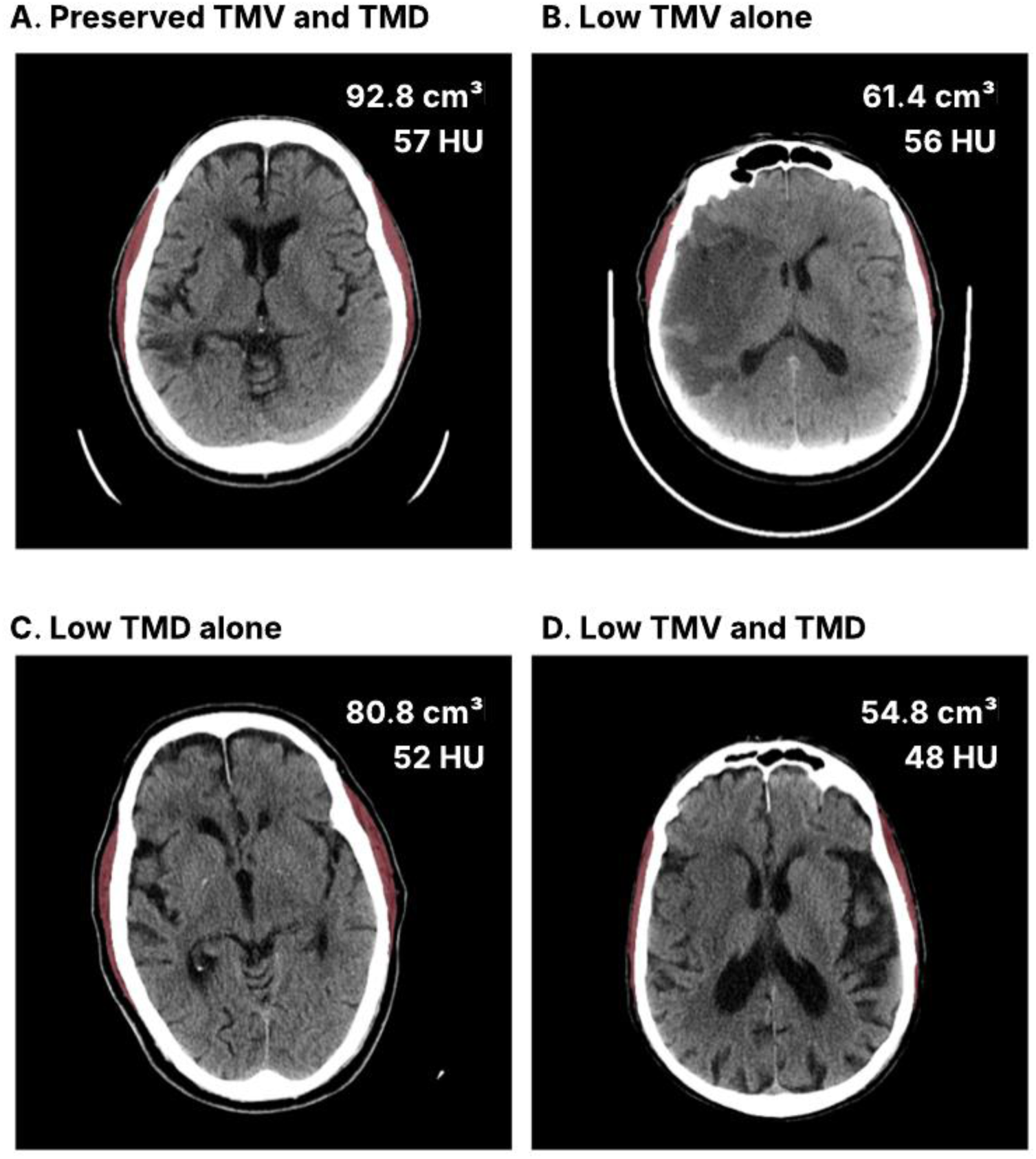
Temporal muscle sarcopenia status by CT. Representative axial head CT images illustrating temporal muscle sarcopenia phenotypes defined by TMV and TMD. (A) Preserved TMV and TMD. (B) Isolated low TMV with preserved TMD. (C) Isolated low TMD with preserved TMV. (D) Combined low TMV and low TMD. All patients shown were male given the sex-specific TMV thresholds. Bilateral temporal muscles are segmented and overlaid (red). TMV is reported in cm³ and TMD in HU. Abbreviations: CT, computed tomography; TMV, temporal muscle volume; TMD, temporal muscle density; HU, Hounsfield units.

Table 1 presents baseline characteristics stratified by sarcopenia status. 877 patients (38%) were non-sarcopenic, 838 (37%) were pre-sarcopenic, and 570 (25%) were sarcopenic. Sarcopenic patients were older, more often female, had lower body weight and higher comorbidity burden, and were more likely to arrive by ambulance. Patients classified as pre-sarcopenic were intermediate for most characteristics.

**Table 1.** Baseline characteristics by sarcopenia status.

|  | No sarcopenia<br>N=877 | Pre-sarcopenia<br>N=838 | Sarcopenia<br>N=570 | *P* value |
| --- | --- | --- | --- | --- |
| Age, mean (SD), y | 67.8 (14.4) | 78.1 (11.8) | 80.4 (10.1) | < 0.001 |
| Female sex, No. (%) | 365 (42) | 532 (63) | 254 (45) | < 0.001 |
| Height, mean (SD), cm | 168.3 (9.9) | 164.3 (9.5) | 168.2 (9.6) | < 0.001 |
| Weight, mean (SD), kg | 75.9 (17.3) | 70.3 (17.0) | 71.2 (16.9) | < 0.001 |
| BMI, mean (SD), kg/m<sup>2</sup> | 27.0 (5.2) | 26.5 (5.8) | 26.3 (4.7) | 0.120 |
| Arrival by ambulance | 468 (57) | 540 (71) | 388 (77) | < 0.001 |
| Arrival after hours | 430 (49%) | 417 (50%) | 279 (49%) | 0.940 |
| HFRS, mean (SD) | 6.5 (5.3) | 9.1 (6.8) | 10.0 (7.4) | < 0.001 |
| Atrial fibrillation | 164 (19) | 268 (32) | 201 (35) | < 0.001 |
| Congestive heart failure | 57 (6) | 114 (14) | 95 (17) | < 0.001 |
| Coronary artery disease | 162 (18) | 182 (22) | 169 (30) | < 0.001 |
| Peripheral artery disease | 66 (8) | 78 (9) | 60 (11) | 0.131 |
| Chronic kidney disease | 213 (24) | 276 (33) | 213 (37) | < 0.001 |
| Diabetes mellitus | 316 (36) | 304 (36) | 199 (35) | 0.862 |
| Hypertension | 642 (73) | 689 (82) | 453 (79) | < 0.001 |
| Dyslipidemia | 414 (47) | 429 (51) | 308 (54) | 0.033 |
| Dementia | 57 (6) | 122 (15) | 103 (18) | < 0.001 |
| Malnutrition | 39 (4) | 64 (8) | 55 (10) | < 0.001 |
| Anemia | 121 (14) | 177 (21) | 153 (27) | < 0.001 |

Baseline demographic, anthropometric, and clinical characteristics of the study cohort stratified by sarcopenia status. Continuous variables are presented as mean (SD), and categorical variables as number (%). Group comparisons were performed using linear regression tests for continuous variables and $\chi^2$ tests for categorical variables, as appropriate. *P* values reflect overall comparisons across sarcopenia groups.
Abbreviations: BMI, Body Mass Index; HFRS, Hospital Frailty Risk Score; SD, Standard Deviation.

### Association of CT-derived muscle metrics with 30-day mortality

30-day mortality was 22 (3%), 66 (8%), and 51 (9%) in the non-sarcopenic, pre-sarcopenic, and sarcopenic patients (*P* < 0.001). Adjusted odds ratios (OR) were 2.70 (95% CI 1.64 to 4.46) and 2.91 (95% CI 1.72 to 4.91) for pre-sarcopenia and sarcopenia (Table 2). When evaluated as continuous metrics, each 1-SD decrease in TMV and TMD was associated with a 38%-41% increase in 30-day mortality odds (TMV_Male_ OR 1.38, 95% CI 1.04 to 1.83; TMV_Female_ OR 1.41, 95% CI 1.07 to 1.84; TMD OR 1.41, 95% CI 1.12 to 1.77) (Table S1). The AUROC of the baseline model increased from 0.676 to 0.722 after adding TMV and TMD (*P* = 0.006) (Figure S4A). Indexation of TMV (TMVi) for skull volume yielded similar results. Decreasing TMVi was associated with progressively increasing odds of adjusted 30-day mortality irrespective of TMD (Figure 2).

**Figure 2.**
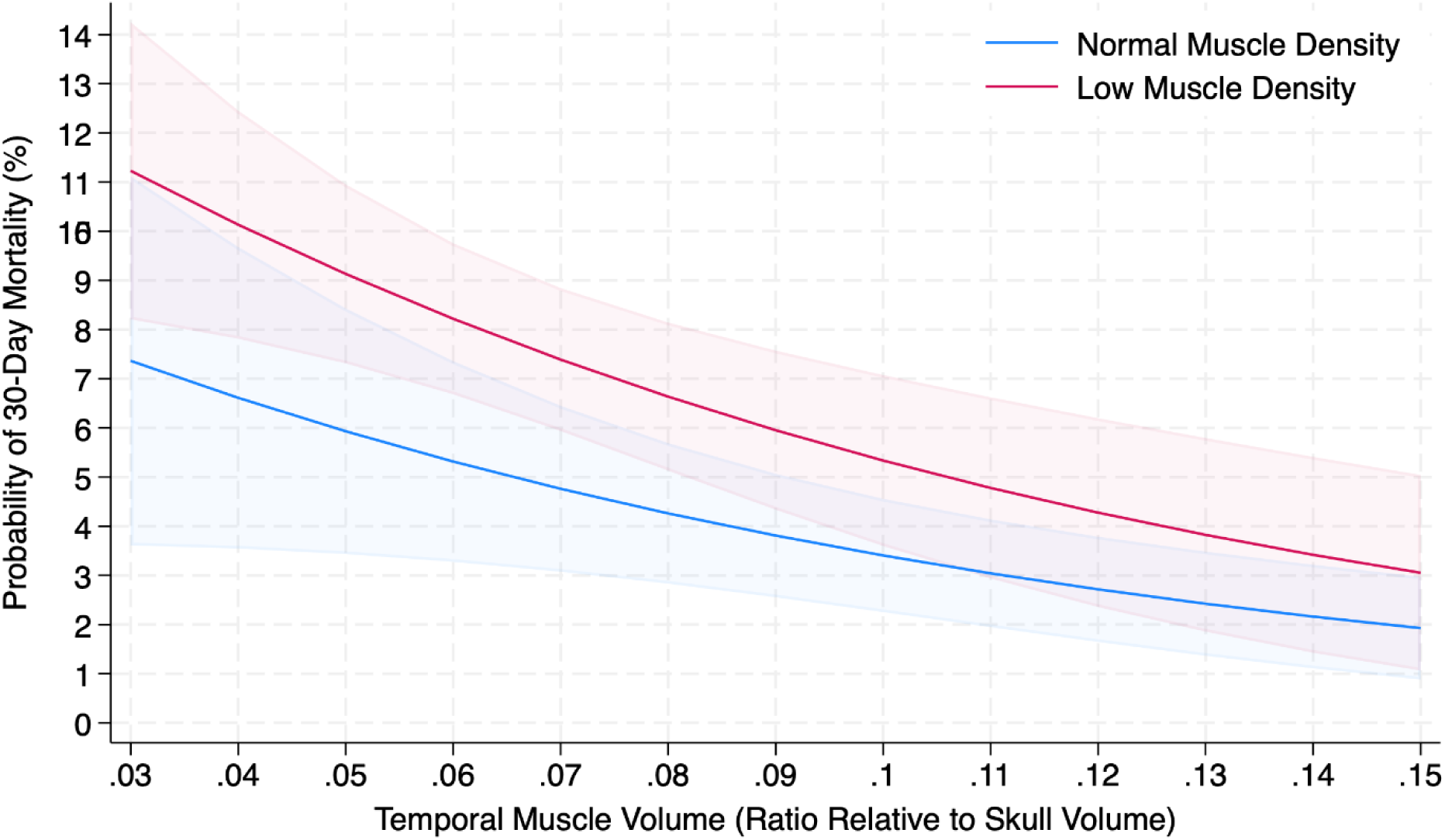
Adjusted margins for 30-day mortality by TMVi. Adjusted marginal probabilities of 30-day all-cause mortality plotted across the range of TMVi. Curves are stratified by TMD, comparing normal muscle density and low muscle density. Shaded bands represent 95% CIs. Decreasing TMVi is associated with progressively higher adjusted mortality risk across the full range of TMD, indicating that the association between lower TMVi and 30-day mortality is independent of muscle density. Abbreviations: TMVi, indexation of temporal muscle volume for skull volume; TMD, temporal muscle density; CI, confidence interval.

**Table 2.** Multivariable models for 30- and 365-day mortality by sarcopenia status.

|  | **30-Day Mortality** |  | **365-Day Mortality** |  |
| --- | --- | --- | --- | --- |
|  | *OR (95% CI)* | *P value* | *OR (95% CI)* | *P value* |
| **Without HFRS** |  |  |  |  |
| Predicted Mortality | 1.13 (1.09 to 1.18) | < 0.001 | 1.15 (1.11 to 1.18) | < 0.001 |
| Sarcopenia Status |  |  |  |  |
| Pre-sarcopenia | 2.70 (1.64 to 4.46) | < 0.001 | 2.42 (1.74 to 3.36) | < 0.001 |
| Sarcopenia | 2.91 (1.72 to 4.91) | < 0.001 | 2.96 (2.09 to 4.17) | < 0.001 |
| AUROC | 0.705 |  | 0.714 |  |
| **With HFRS** |  |  |  |  |
| Predicted Mortality | 1.11 (1.06 to 1.16) | < 0.001 | 1.12 (1.08 to 1.15) | < 0.001 |
| HFRS Class |  |  |  |  |
| Intermediate | 3.10 (1.79 to 5.36) | < 0.001 | 3.00 (2.05 to 4.40) | < 0.001 |
| High | 3.64 (1.96 to 6.76) | < 0.001 | 7.57 (4.97 to 11.51) | < 0.001 |
| Sarcopenia Status |  |  |  |  |
| Pre-sarcopenia | 2.46 (1.49 to 4.07) | < 0.001 | 2.10 (1.50 to 2.95) | < 0.001 |
| Sarcopenia | 2.61 (1.54 to 4.43) | < 0.001 | 2.41 (1.69 to 3.43) | < 0.001 |
| AUROC | 0.735 |  | 0.770 |  |

Multivariable logistic regression models evaluating the association between sarcopenia status and 30-day and 365-day all-cause mortality. Models are presented without and with adjustment for HFRS. ORs are reported with 95% CIs. Model discrimination is summarized using the AUROC curve.
Abbreviations: HFRS, Hospital Frailty Risk Score; AUROC, Area Under the Receiver Operating Characteristic; OR, odds ratio; CI, confidence interval.

### Association of CT-derived Muscle Metrics with 365-day Mortality

365-day mortality was 57 (6%), 144 (17%), and 121 (21%) in the non-sarcopenic, pre-sarcopenic, and sarcopenic patients (*P* < 0.001). Kaplan-Meier curves are shown in Figure S5. Adjusted ORs were 2.42 (95% CI 1.74 to 3.36) and 2.96 (95% CI 2.09 to 4.17) for pre-sarcopenia and sarcopenia (Table 2). When evaluated as continuous metrics, each 1-SD decrease in TMV and TMD was associated with a 30%-43% increase in 365-day mortality odds (TMV_Male_ OR 1.39, 95% CI 1.14 to 1.69; TMV_Female_ OR 1.30, 95% CI 1.08 to 1.56; TMD OR 1.43, 95% CI 1.22 to 1.69) (Table S1). The AUROC of the baseline model increased from 0.686 to 0.737 after adding TMV and TMD (*P* < 0.001) (Figure S4B).

### Association of CT-derived Muscle Metrics with LOS

The mean (SD) LOS was 13.3 (14.9) days, 19.5 (23.5) days, and 21.1 (24.9) days in the non-sarcopenic, pre-sarcopenic, and sarcopenic patients (*P* < 0.001). Adjusted additional days in hospital were 2.79 (95% CI 1.69 to 3.98) and 3.26 (95% CI 2.00 to 4.64) for pre-sarcopenia and sarcopenia (Table S2). When evaluated as continuous metrics, each 1-SD decrease in TMV and TMD was associated with a 0.65-0.84 increase in days in hospital (TMV_Male_ 0.79, 95% CI 0.24 to 1.37; TMV_Female_ 0.65, 95% CI 0.12 to 1.20; TMD 0.84, 95% CI 0.29 to 1.42) (Table S3).

### Hospital Frailty Risk Score

The mean (SD) HFRS was 6.5 (5.3), 9.1 (6.8), and 10.0 (7.4) in the non-sarcopenic, pre-sarcopenic, and sarcopenic patients (*P* < 0.001). The proportion of patients with low, intermediate, and high-risk HFRS by sarcopenia status is shown in Figure 3. The association between CT-derived sarcopenia and mortality was preserved after adding HFRS to regression models (Tables 2 and S1), with some attenuation for LOS only (Tables S2 and S3). Final regression models for 30- and 365-day mortality, consisting of TMV, TMD, HFRS, and predicted mortality, achieved AUROC of 0.743 and 0.790 (Figure S6).

**Figure 3.**
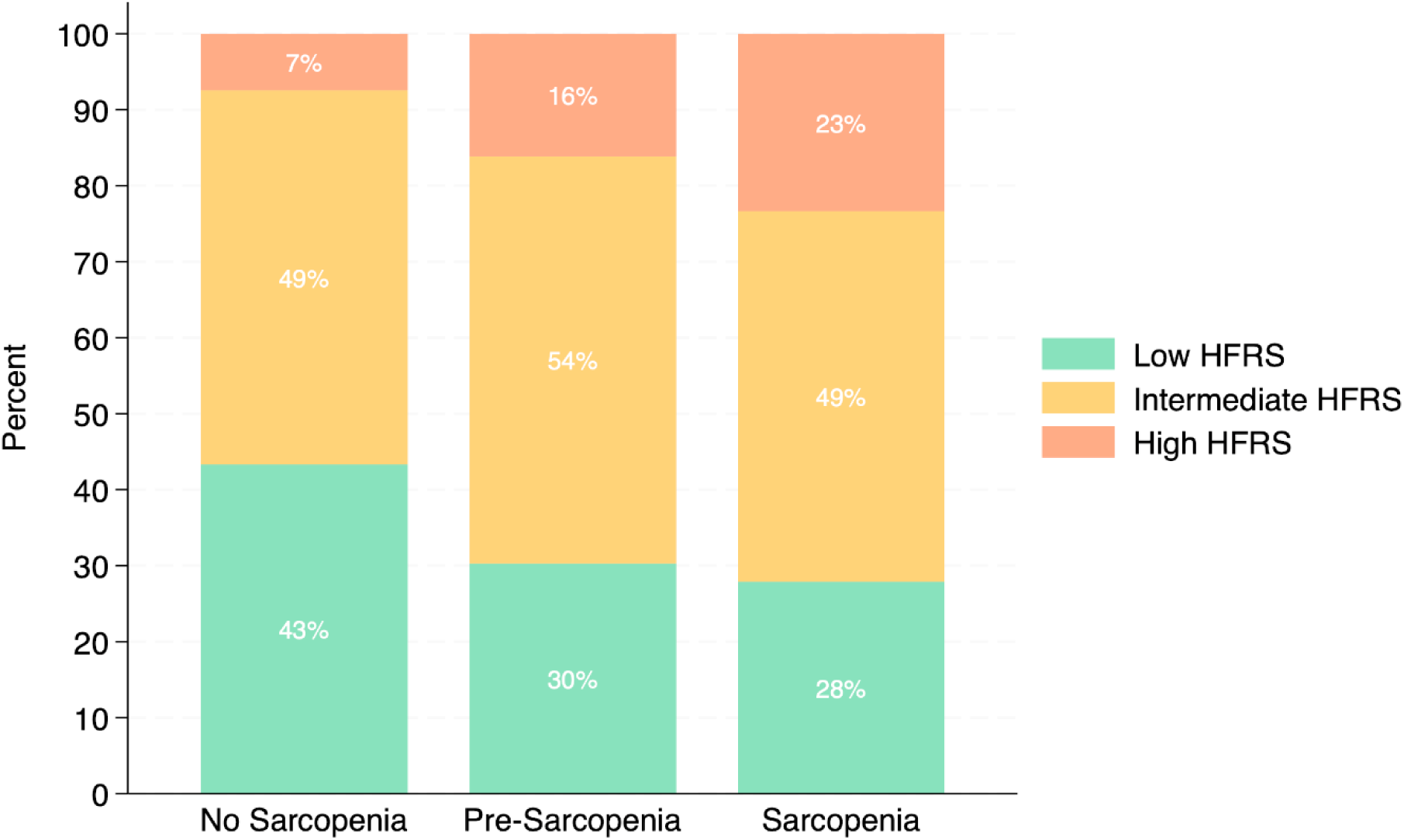
Distribution of HFRS by sarcopenia status. Stacked bar plots showing the proportion of patients classified as low, intermediate, or high HFRS by sarcopenia status. Increasing sarcopenia severity is associated with a graded shift toward higher HFRS categories, with a lower proportion of low-risk and a higher proportion of high-risk HFRS classifications among sarcopenic patients. Abbreviations: HFRS, Hospital Frailty Risk Score.

## DISCUSSION

This study demonstrates that, in AIS patients, routine CT scans can be leveraged using DL to extract novel sarcopenia metrics that incrementally help predict fatal outcomes. The main finding is that the combination of low TMV and TMD predicted a 3-fold increase in short- and mid-term mortality following AIS, as well as prolonged LOS. This association was additive to existing clinical and frailty risk scores. CT-derived sarcopenia was correlated and complementary with the comorbidity-based measure of frailty (HFRS), offering new insight into another dimension of patient frailty. While previous studies have explored manually measured TMT, which has low reliability, this study leveraged a DL-automated and highly accurate method to generate three-dimensional TMV and, importantly, TMD.

Temporal muscle has emerged as a promising, opportunistic biomarker of sarcopenia.^24,25^ Initial studies evaluated manually-segmented TMT and TMA as predictors of postoperative risk and all-cause mortality in patients with hemorrhagic stroke,^16,17,26–28^ glioblastoma,^29–33^ and metastatic brain tumour.^34–36^ Few studies have evaluated TMT or TMA as predictors of AIS prognosis, and among them findings have been inconsistent. This is due in part to the variability of approaches used to quantify the temporal muscle,^11^ selective inclusion criteria focusing on specific stroke subtypes or reperfusion therapies,^14,15^ and relatively small sample sizes used to assess the relationship between temporal muscle characteristics and AIS prognosis.^14,15,37,38^

### Temporal Muscle Quantification in AIS

Ravera et al. included 261 AIS patients treated with reperfusion therapy and determined that lower TMT on admission CT was independently associated with a 1.42-fold increase 90-day mortality odds.^15^ However, they could not establish an independent association between TMT and their primary outcome: 90-day functional status. Nozoe et al. also found no association between TMT and 90-day function status among 289 patients.^19^ Dubinski et al. included 282 cerebellar ischemic stroke patients, and determined that lower TMT on admission CT was independently associated with a 3.4-fold decrease in odds of 365-day poor functional status.^14^

Our study addresses key methodological limitations of these studies. First, our sample size is approximately 10-fold, and thus adequately powered and more representative of diverse AIS subgroups, including patients who are managed supportively.^39,40^ Second, our DL approach automates the process of volumetrically quantifying temporal muscle size. It is therefore not subject to the same measurement variability whereby TMT has previously been measured above, below, or at the level of the orbital roof and Sylvian fissure. Third, our approach yields TMD, which had yet to be investigated and was found to be more strongly associated than TMV with our outcomes.

In the field of MRI, Li et al. included 265 AIS patients, and determined that lower TMT on admission MRI was independently associated with a 3.5-fold increase in mortality hazard and shorter survival.^38^ Huang et al. included 467 AIS patients, and determined that lower TMT and TMA on admission MRI using an end-to-end DL pipeline, were associated with 180-day poor functional status.^37^ The translation of these findings to acute care is limited by their reliance on MRI, since CT is the imaging modality most frequently used at the acute point of care.^41^

### Impact of Muscle Volume and Density

TMD added incremental value over TMV alone, and allowed for a trichotomous classification of patients (non-sarcopenic, pre-sarcopenic, sarcopenic) similar to European Working Group for Sarcopenia in Older People (EWGSOP) guidelines.^8^ Pre-sarcopenic patients generally showed intermediate features between non-sarcopenic and sarcopenic patients across baseline characteristics and outcomes. Of the 838 pre-sarcopenic patients, 636 had low TMD with preserved TMV, and therefore would have been misclassified as non-sarcopenic based on TMV alone. These novel findings underscore the importance of measuring both volume and density to assess sarcopenia adequately.

### Temporal Muscle Size, Sarcopenia, and Frailty

Multiple independent cohorts now link smaller temporal muscle size to both sarcopenia and clinical frailty. Narrative and systematic reviews across stroke and neuro-oncology populations established TMT as a correlate of skeletal muscle mass, functional capacity, and nutritional status.^11,25,42–44^ Two prospective studies, conducted in AIS patients, established TMT as a valid surrogate of sarcopenia as diagnosed by the SARC-F instrument^19^ or bioimpedance analysis with handgrip strength.^45^ Another study, conducted in 500 healthy adults, confirmed TMT as a valid surrogate of sarcopenia as diagnosed by EWGSOP criteria.^20^ Together, these data validate temporal muscle metrics as imaging biomarkers of skeletal muscle mass and sarcopenia.

### Clinical Relevance of TMV and TMD in Stroke Care

DL-derived TMV and TMD from clinical CT scans address an unmet need for geriatric risk stratification in AIS. Routine integration of these metrics may be used to refine risk estimates, guide informed, shared decision-making (including life-sustaining therapies), and personalize therapeutic plans to optimize patient-centred outcomes. One exciting albeit unproven application may be to triage patients for aggressive therapies such as endovascular treatment or thrombolysis, which have muted benefits and higher risks of iatrogenic complications in the presence of frailty. For example, AIS patients with clinical frailty have been shown to be less likely to respond favorably to endovascular treatment ^46^ and thrombolysis.^47^ Prospective clinical trials are eagerly awaited to test the hypothesis of frailty-informed clinical care pathways for AIS.

Once low TMV or TMD is identified, a first action may be to test for reversible causes of sarcopenia, such as protein-energy malnutrition or androgen deficiency, and treat accordingly. Protein-enriched dietary supplements have been shown to stimulate muscle synthesis and prevent incremental losses during the catabolic phase of an acute hospitalization.^48^ Another action may be to prioritize these patients for referral to a dietician and speech language pathologist, as dietary intake and swallowing capabilities are likely reduced,^49,50^ and to anticipate referral to a rehabilitation centre, as temporal sarcopenia is likely to be associated with generalized sarcopenia and weakness.^19,20,45^ Frail patients may also benefit from referral – either during the acute hospitalization or thereafter – to a geriatric specialist for comprehensive geriatric assessment and intervention.

### Limitations

The first limitation is this study’s single-centre nature, limiting its generalizability to outside centres. However, this study included multiple CT scanners from different vendors, and a large multi-ethnic patient sample. The second limitation is the lack of baseline or follow-up NIHSS scores and adjudicated functional outcomes, clearly relevant for older patients suffering from frailty. Future research on TMV and TMD should determine the extent to which these metrics modulate functional recovery and physical performance post-AIS. The third limitation is the paucity of normative reference data for TMV and TMD, which was partially addressed in this study by collecting CT scans in a sample of healthy adults and by representing TMV and TMD as continuous variables rather than as binary classes using thresholds.

## CONCLUSIONS

In AIS patients, TMV and TMD were strongly predictive of mortality and were readily extracted from clinical CT scans using a novel, DL-automated pipeline. This pipeline is scalable, rapid, and does not require manual input from healthcare professionals. TMV and TMD added incremental value to traditional clinical risk and frailty scores, providing insight into sarcopenia, a distinct facet of frailty. Incorporating these metrics into standard workflows would enhance risk stratification and catalyze frailty-focused interventions with potential to improve patient-centred outcomes.

## Data Availability

Requests for data sharing will be considered on a case-by-case basis following institutional governance policies.

## ACKNOWLEDGMENTS

The authors have no acknowledgments to declare.

## SOURCES OF FUNDING

Joshua Ron Onichino, MD(c), is supported the *Dr. Clarke K. McLeod Memorial Scholarship* and the *Class of Medicine 1979 Research Bursary* issued by the McGill University Faculty of Medicine and Health Sciences.

Dr. Jonathan Afilalo, MD, MSc, FACC, is supported by research grants issued by the *Fonds de recherche du Québec - Santé* and the Canadian Institutes of Health Research.

## DISCLOSURES

Dr. Jonathan Afilalo, MD, MSc, FACC, is the creator of CoreSlicer, an open-access platform for research on body composition analysis using clinical imaging scans.

## NON-STANDARD ABBREVIATIONS AND ACRONYMS

TMV: temporal muscle volume
TMD: temporal muscle density
TMT: temporal muscle thickness
TMA: temporal muscle area
TMVi: indexation of TMV for skull volume
HFRS: Hospital Frailty Risk Score
LOS: length of stay

## Notes

### Competing Interest Statement

The authors have declared no competing interest.

### Author Declarations

The study was approved by the CIUSSS West-Central Montreal Research Ethics Board, which granted a waiver of individual informed consent for secondary analyses of existing clinical and imaging data.

